# Comparison of SARS-COV-2 nasal antigen test to nasopharyngeal RT-PCR in mildly symptomatic patients

**DOI:** 10.1101/2020.11.10.20228973

**Authors:** Abdulkarim Abdulrahman, Fathi Mustafa, Abdulla I AlAwadhi, Qadar Alansari, Batool AlAlawi, Manaf AlQahtani

**Affiliations:** National Taskforce for Combating the Coronavirus (COVID-19), Bahrain; Mohammed Bin Khalifa Cardiac Centre, Bahrain; Royal College of Surgeons in Ireland, Bahrain; Bahrain Defence Force hospital, Bahrain; Ministry of Finance and National Economy, Bahrain; Ministry of Health, Bahrain

## Abstract

**Introduction:** COVID 19 has been vastly spreading since December 2019 and the medical teams worldwide are doing their best to limit its spread. In the absence of a vaccine the best way to fight it is by detecting infected cases early and isolate them to prevent its spread. Therefore, a readily available, rapid, and cost-effective test with high specificity and sensitivity for early detection of COVID 19 is required. In this study, we are testing the diagnostic performance of a rapid antigen detection test in mildly symptomatic cases. (RADT).

**Methods:** The study included 4183 patients who were mildly symptomatic. A nasal sample for the rapid antigen test and a nasopharyngeal sample was taken from each patient. Statistical analysis was conducted to calculate the sensitivity, specificity, positive predictive value, negative predictive value and kappa coefficient of agreement.

**Results:** The prevalence of COVID 19 in the study population was 17.5% (733/4183). The calculated sensitivity and specificity were 82.1% and 99.1% respectively. Kappa’s coefficient of agreement between the rapid antigen test and RT-PCR was 0.859 (p < 0.001). A stratified analysis was performed and it showed that the sensitivity of the test improved significantly with lowering the cutoff Ct value to 24.

**Conclusion:** The results of the diagnostic assessment of nasal swabs in the RADT used in our study are promising regarding the potential benefit of using them as a screening tool in mildly symptomatic patients. The diagnostic ability was especially high in cases with high viral load. The rapid antigen test is intended to be used alongside RT-PCR and not replace it. RADT can be of benefit in reducing the use of PCR.

## Introduction

Since December 2019, the number of Coronavirus disease (COVID 19) confirmed cases has been rising swiftly despite the efforts to limit its spread. ^1, 2^ The World Health Organization declared COVID 19 to be a pandemic on 12^th^ March 2020.^2^ To date, the total number of cases and deaths worldwide has exceeded 50 million and 1 million, respectively.^3^ The medical team in Bahrain has been working diligently to confine the spread of this disease since the pandemic started. Bahrain have had more than 83000 COVID19 cases, with about of 5.56% of the population affected.^4^

The best way to battle this virus in the absence of an effective vaccine is the ability to detect infections early enough to take the necessary precautions to halt its spread to contacts and adequately manage high-risk patients. What is challenging about that is the lack of a readily available, rapid, and cost-effective test with high specificity and sensitivity for early detection of COVID 19 infected patients in the general population.^5-8^

Until now, nasopharyngeal Real Time – Polymerase Chain Reaction (RT-PCR) is the gold standard diagnostic test for COVID 19.^5-8^ RT-PCR has multiple limitations such as delayed results availability and the need for specialized laboratory equipment as well as specialized technicians.^1, 5, 6, 8^ As a result, the number of tests performed per day is limited, and the appropriate management of positive cases might be delayed. Therefore, other diagnostic techniques are needed to limit the spread of the virus and effectively monitor the degree of infection in COVID 19 patients.^1, 5, 6, 8^ Current literature explored the possibility of using point-of-care rapid antigen test as it is cost-effective, simple and was used effectively with other viruses such as Influenza and RSV.^9^ However, the studies showed that it had low overall sensitivity and high specificity compared to RT-PCR.^1, 5, 6, 8^

In our study, we are exploring the diagnostic performance of nasal swabs as they do not require a skilled professional in addition to being less time consuming and cause less discomfort. We aim to demonstrate the efficacy of nasal antigen tests in mildly symptomatic cases. This would provide a simple reliable test that might eliminate negative cases with a certain level of confidence. In addition, implementation of such tests will reduce the workload on healthcare professionals, as these tests can be done at clinics or home, and ease the process of reopening.

### Objective

Determine the accuracy of the nasal swab antigen test in the detection of SARS-COV-2 compared to nasopharyngeal RT-PCR in mildly symptomatic individuals.

## Methods

### Study population

The study involved 4183 mild symptomatic individuals. Mild symptomatic individuals definition followed Bahrain protocol^10^. It included symptoms such as fever (<38°C), loss of taste or smell, flu-like symptoms, sore throat, GI symptoms, myalgias and fatigue. The study participants were referred to the symptomatic hall in the national testing centre at the exhibition center.

### Setting

All testing was conducted in the symptomatic hall in the National Testing Centre at the Exhibition centre

### Study design

We conducted a cross-sectional study to determine the diagnostic performance of the rapid antigen test compared to RT-PCR. Two swabs were taken from each person, one nasal swab for the antigen test and one nasopharyngeal swab for the RT-PCR. For rapid antigen test, Abbott panbio COVID 19 antigen rapid test device to detect SARS-CoV-2 nucleocapsid protein was used (Abbott Rapid Diagnostic Jena GmbH, Jena, Germany) ^11^. The nasopharyngeal samples for RT-PCR were transferred to a viral transport media immediately after collection and transported to a COVID-19 laboratory for testing. The PCR test was conducted using Thermo Fisher Scientific (Waltham, MA) TaqPath 1-Step RT-qPCR Master Mix, CG on the Applied Biosystems (Foster City, CA) 7500 Fast Dx RealTime PCR Instrument. The assay used followed the WHO protocol and targeted the E gene. If the E gene was detected, the sample was then confirmed by RdRP and N genes ^12^. The E gene Ct value was reported and used in this study. Ct values >40 were considered negative. Positive and negative controls were included for quality control purposes.

### Sample collection

All samples were collected by a trained healthcare professional in the National testing centre. The nasal samples were collected, using the nasopharyngeal swab provided with the RADT kit from both nostrils. Based on the CDC guidelines, the patient’s head was tilted laid back by 70°, then the swab was inserted by approximately 2cm into the nostril while gently rotating it, rolled it several times then removed it.

The nasopharyngeal samples used for PCR were collected through both nostrils from the nasopharynx using a nasopharyngeal swab ^13^. The nasopharyngeal swab was inserted into the nostril parallel to the palate until resistance was encountered or the depth was equivalent to the distance of the nose from the ear. The swab was rolled and rubbed gently, left in place for multiple seconds, then removed slowly while rotating it ^13^.

### Participants

- Inclusion criteria
  - suspected COVID 19 cases with mild symptoms (defined by Bahrain protocol ^10^) presenting to the testing centre
- Exclusion criteria
  - Suspected cases with severe symptoms
  - Any asymptomatic suspected case

### Data Handling and statistical analysis

Antigen test results and RT-PCR result with the corresponding Ct value were collected for all mildly symptomatic cases. The diagnostic performance of the antigen was assessed using sensitivity, specificity, positive predictive value, negative predictive value and their respective 95% Confidence interval. Agreement between nasopharyngeal PCR and nasal antigen tests was assessed using kappa coefficient of agreement. The Ct value of identified and missed cases by antigen tests were summarized using median and interquartile range. Ct Value of identified and missed cases were compared using a two sample T test. All P-values were two-sided and P < 0.05 was considered significant. Data collection was performed through a live google sheet and extracted to Microsoft Excel 2016. Statistical analysis was performed using STATA (StataCorp. 2017)

### Ethical considerations

Ethical and research approval was obtained from the National COVID19 research and ethics committee (approval code: CRT-COVID-2020-088). All methods and analysis of data was approved by the National COVID-19 Research and Ethics Committee and carried out in accordance with the local guideline and ethical guidelines of the Declaration of Helsinki 1975. Written Informed consent was waived by the Research and Ethical Committee for this study due to the absence of any patient identifying information.

## Results

A total of 4183 mild symptomatic cases were tested by PCR (using a nasopharyngeal sample) and by antigen test (using a nasal sample). 56.5% of the cases were males and 43.5% were females. Mean age of the tested population was 30.9yrs (± 14.5yrs). Days from symptom onset ranged from 0 to 14 with a median of 2 (IQR 1 - 3). Table1 summarizes the demographics of the tested cohort. 17.5% (733/4183) of the population tested positive by PCR, no equivocal results were reported. Using the antigen test, 15.1% were positive, while the remaining tested negative and none of the tests were equivocal.

**Table 1.** Demographics and clinical features of studied sample.

| Variables | N | Value |
| --- | --- | --- |
| Age in years – Mean ± SD | 4183 | 30.9 ± 14.5 |
| Male – no. (%) | 4183 | 2365 (56.5%) |
| Symptoms Onset in days - Median (IQR) | 1301 | 2 (1- 3) |
| Prevalence – no. (%) | 4183 | 733 (17.5%) |
| Ct Value of PCR positive cases - Median (IQR) | 530 | 22 (20 – 24.5) |

Out of the 733 confirmed cases by PCR, the antigen test accurately diagnosed 602 cases (82.1%). 135 cases were falsely negative by the antigen test and 30 cases were reported as false positive. Table 2 is a contingency table showing the PCR and Antigen test results. Using nasopharyngeal RT-PCR as the gold standard test for diagnosis of SARS-CoV2, the rapid antigen test showed a sensitivity of 82.1% (95% CI 79.2% - 84.8%) and a specificity of 99.1% (95% CI 98.8% - 99.4%). With the prevalence of COVID19 being 17.5% within the tested population, the antigen test had a Positive predictive value (PPV) of 95.3% and a Negative predictive value (NPV) of 96.3%. Agreement analysis between the nasopharyngeal PCR and the nasal antigen test showed 85.9% observed agreement (κ coefficient = 0.859, p< 0.001). Table 3 summarizes the diagnostic performance of the antigen test.

**Table 2:** 2×2 table showing the PCR and Antigen test results.

|  | PCR Positive | PCR Negative | Total Antigen Results |
| --- | --- | --- | --- |
| Antigen Test Positive | 602<br>True Positive | 30<br>False Positive | 632 |
| Antigen Test Negative | 131<br>False Negative | 3420<br>True Negative | 3551 |
| Total PCR Results | 733 | 3450 | Total Cases: 4183 |

**Table 3:** Assessment of the diagnostic accuracy of the antigen test.

|  | Value | 95% CI |  |
| --- | --- | --- | --- |
| Prevalence | 17.5% | 16% | 18.7% |
| Sensitivity | 82.1% | 79.2% | 84.8% |
| Specificity | 99.1% | 98.8% | 99.4% |
| Positive Predicted Value | 95.3% | 93.3% | 96.8% |
| Negative Predicted Value | 96.3% | 95.6% | 96.9% |
| Positive Likelihood Ratio | 94.4 | 66 | 135 |
| Negative Likelihood Ratio | 0.18 | 0.154 | 0.211 |
| False Discovery Rate | 3.8% | 3.3% | 4.4% |
| Kappa Coefficient | 85.9% | 83.8% to 88% (p < 0.001) |  |

Confirmed cases had a median Ct value of 22 (IQR 20 - 24.1). Cases detected by antigen test had a median Ct value of 22 (IQR 2-to 24) and a mean of 22.1 (95% CI 21.9 - 22.4) Cases missed by the antigen test had a median Ct value of 25 (IQR 22 – 28) and a mean of 25.1 (95% CI 24.3 - 25.8). Mean Ct value difference between the false negative and the true positive cases was statistically significant (t-score 9.2, p<0.001). The median Ct values and their corresponding interquartile ranges are shown in figure 1.

**Figure 1.**
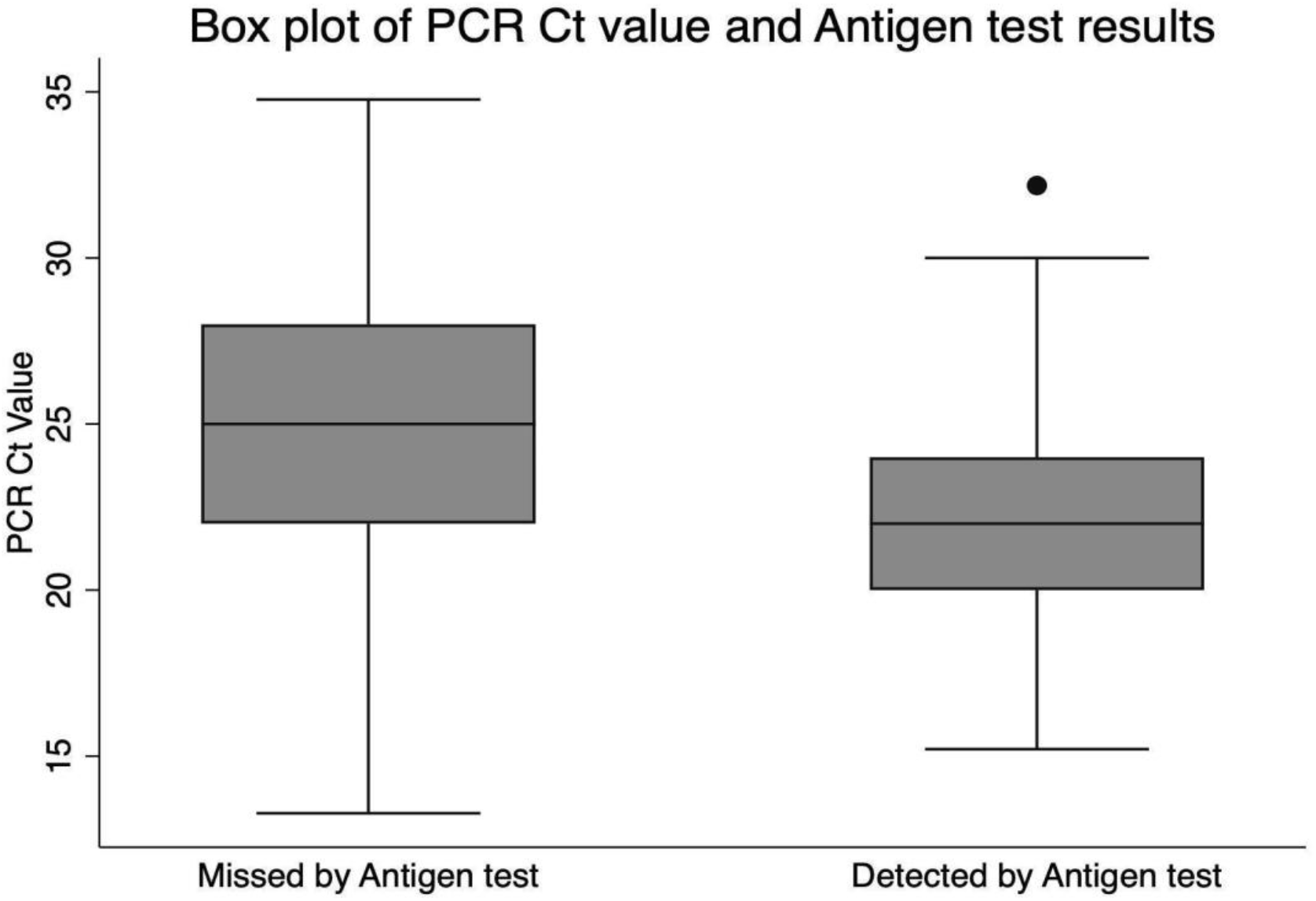
Comparison of the median Ct values between detected and missed cases by the antigen test.

To control for time since symptom onset as a confounder, we performed a stratified analysis to assess the significance of time since onset of symptoms on the diagnostic performance of the antigen test. Cases with symptom onset within 5 days showed a modest improvement in the diagnostic performance with sensitivity of 82.4%, specificity of 99.3% and a Kappa coefficient of 0.865. This was almost similar to cases that had symptom onset within 7 days as shown in Table 4. Additionally, a secondary analysis was conducted after excluding cases with Ct value more than or equal to 30 and Ct more than 24. The sensitivity increased to 84.5% and 87.9% respectively. Whereas specificity for both cutoff Ct values was 99.1%. Moreover, after excluding cases with Ct value > 24 and restricting symptoms onset to within 5 days and 7 days, there was a significant increase in sensitivities up to 89.5% and 89.3% respectively.

**Table 4:** The effect of symptoms onset time and Ct values on the diagnostic performance.

|  | Model | N | Prevalence | Sensitivity | Specificity | Negative Predicted Value | Positive Predicted Value |
| --- | --- | --- | --- | --- | --- | --- | --- |
| 1 | Symptom onset within 7 days | 1290 | 20%<br>(18% - 22.8%) | 82.6%<br>(77.5% - 87%) | 99.3%<br>(98.6% - 99.7%) | 95.7%<br>(94.3% - 96.8%) | 96.9%<br>(93.7% - 98.7%) |
| 2 | Symptom onset within 5 days | 1252 | 20%<br>(18% - 22.8%) | 82.4%<br>(77.2% - 86.9%) | 99.3%<br>(98.6% - 99.7%) | 95.6%<br>(94.2% - 96.8%) | 96.8%<br>(93.5% - 98.7%) |
| 3 | Excluding cases with Ct ≥ 30 | 4148 | 17%<br>(16% - 18.1%) | 84.5%<br>(81.6% - 87.1%) | 99.1%<br>(98.8% - 99.4%) | 96.9%<br>(96.3% - 97.5%) | 95.2%<br>(93.2% - 96.7%) |
| 4 | Excluding cases with Ct > 24 | 3996 | 14%<br>(13% - 15%) | 87.9%<br>(84.9% - 90.5%) | 99.1%<br>(98.8% - 99.4%) | 98.1%<br>(97.6% - 98.5%) | 94.2%<br>(91.8% - 96.1%) |
| 5 | Symptom onset within 7 days and Excluding cases with Ct ≥ 30 | 1274 | 20%<br>(17% - 21.8%) | 86.3%<br>(81.4% - 90.4%) | 99.3%<br>(98.6% - 99.7%) | 96.8%<br>(95.5% - 97.8%) | 96.8%<br>(93.6% - 98.7%) |
| 6 | Symptom onset within 7 days and Excluding cases with Ct > 24 | 1220 | 16%<br>(14% - 18.3%) | 89.3%<br>(84.2% - 93.3%) | 99.3%<br>(98.6% - 99.7%) | 98%<br>(96.9% - 98.7%) | 96.2%<br>(92.3% - 98.4%) |
| 7 | Symptom onset within 5 days and Excluding cases with Ct > 30 | 1236 | 19%<br>(17% - 21.8%) | 86.3%<br>(81.3% - 90.4%) | 99.3%<br>(98.6% - 99.7%) | 96.8%<br>(95.5% - 97.8%) | 96.7%<br>(93.4% - 98.7%) |
| 8 | Symptom onset within 5 days and Excluding cases with Ct > 24 | 1184 | 16%<br>(14% - 18.3%) | 89.5%<br>(84.2% - 93.5%) | 99.3%<br>(98.6% - 99.7%) | 98%<br>(96.9% - 98.8%) | 96%<br>(92% - 98.4%) |

As a follow up, cases that tested negative by the antigen test and tested positive by the RT-PCR were asked to repeat the antigen test again within 72hrs. 19 out 135 responded and 73.7% were positive on the repeated antigen test. 3 of the 30 cases who tested positive by the antigen test but negative by PCR, were tested again within 72hrs. One case tested negative, while two remained positive and were positive by PCR too.

## Discussion

The PCR has been a very accurate test to diagnose all kinds of infectious diseases. It provides results faster than cultures, and its use for early diagnosis by the infectious disease specialists has been very popular ^14^. During the pandemic, PCR test was the only accurate test available to diagnose infected people ^15^. PCR is a very sensitive test for SARS-CoV2, and this sensitivity had improved within a few months into the pandemic. Some PCR machines detect down to ten viral RNA copies μl–1 ^16^. Despite the PCR test’s high sensitivity, it has multiple limitations that hold back the efforts in battling this pandemic, especially when reopening had started. Multiple studies showed that RT-PCR was sometimes positive in patients with a corresponding negative culture test for SARS-CoV-2, which indicates that these patients were not infectious ^8, 17^. This has led to the isolation of people who are noninfectious, and halted reopening measures. Another limitation is that it requires healthcare professionals to collect the swab and to specialized labs and specialists to analyse and interpret the result ^5, 6^.

As the pandemic necessitated mass testing, the turnaround time extended and required on average 2-3 days in many countries ^18, 19^. This time limitation has kept Bahrain under-armed when fighting the pandemic. One of the main steps forward to fight this pandemic is to have an accurate test that will detect infectious people - who are a public health risk - and return results quickly. The test should also be easily performed by the general population and can be repeated multiple times whenever necessary. This will reduce the workload on healthcare professionals as well as smoothen the reopening process. The use of the nasopharyngeal swab is a limiting factor in the ease and frequency of testing because it is invasive, uncomfortable and aerosolizing ^20^. For similar reasons, the Centers for Disease Control and Prevention (CDC) permitted self-sampling via nasal swabs to compensate for the shortage of healthcare workers and the escalation of COVID 19 cases ^21^. Furthermore, the CDC as well as some studies have illustrated that supervised nasal swabs were quite as effective as nasopharyngeal swabs in detecting SARS-CoV-2 ^14, 22^.

The antigen test used in our study showed that it can be a reasonable test to be used in this context. The nasal antigen test had a significant agreement correlation of 85% with the nasopharyngeal PCR in the studied population. The mild to moderate symptomatic population represent the majority of cases within COVID19, 81% as reported by a Chinese cohort ^23^. Hence targeting this population should be a priority when using a newer test. In cases where patients present with severe disease, PCR test should continue to be used as having a definite result is necessary.

The rapid antigen detection test (RADT) in our study had a very high specificity of 99.3%. The test also had high predictive value within a population with a 18% prevalence of COVID 19. The sensitivity of the test was 82.1% when compared to the PCR test. Despite the antigen test having lower sensitivity, it was done using a nasal and not a nasopharyngeal sample. Moreover the diagnostic accuracy of PCR can never be compared to rapid point of care antigen test as the method of detection is different.

The findings in our study regarding the diagnostic performance of the rapid antigen test match the data in the current literature to a certain extent. For example, a review of nine studies involving 7 different brands of rapid antigen tests reported that all studies reported very high specificities. The pooled specificity was 99% (95% CI 98-100%) which was similar to our test’s specificity (99.2%). However the reported pooled sensitivity was 49% (95% CI 28 - 70) which is completely different from our test’s sensitivity (81.3%). There was a wide range of the sensitivities across the studies from 0 to 94% ^24^. Few high quality studies showed that some tests such as the Bioeasy 2019-nCov Ag Fluorescence Rapid Test Kit had a relatively high pooled sensitivity of 82.3%, which is slightly close to our test’s sensitivity ^25^.

There were multiple studies that reported either low sensitivities such as 30% and 50% or low cohen’s coefficient of agreement while in our study the reported sensitivity was 81.3% ^5, 7, 26^. All of the aforementioned studies were using different commercial antigen tests as well as different swabs (nasopharyngeal swabs and nasal swabs). Moreover, most of the studies did not specify the severity of symptoms within the study population. The studies that reported very high sensitivities of the rapid antigen test usually involved patients who were either in the emergency department or hospitalised. Such patients are usually more symptomatic, hence have a higher viral load. As a result, the reported sensitivities were higher compared to patients with milder symptoms ^27-30^.

The cases missed by the antigen test in our study had higher Ct Value than those detected by the antigen test. The mean Ct value for the missed cases was 25.1. Bullard et al described that viral cultures fail when time from symptom onset exceeds 8 days and/or the ct value exceeds 24 ^31^. When we excluded cases above Ct value of 24, the sensitivity improved to 87.9% with an aggremention rate (Kappa Coefficient) of 89.2 between nasal antigen test and nasopharyngeal PCR. The accuracy improved further when symptom onset was restricted to 7 days and cases above Ct of 24 were excluded. The agreement coefficient reached 91.3% and sensitivity reached 89.3% without affecting specificity. This finding was also reported by Bayona et al in a meta analysis conducted on multiple RADT, the sensitivity of the RADT was higher when performed in patients early in the disease (0 - 7 days) compared to tests performed late in the disease (8 - 14 days). The study also showed that the reduction of the Ct value from ≤ 40 to ≤ 30 increased the sensitivity of the rapid antigen test from 68% to 98%. One of the studies included in the meta-analysis showed that the sensitivity improved to 82.2% in patients with higher viral loads (Ct value < 25)^7^. In addition, the median Ct value of antigen test negative cases was higher and significantly different from positive cases ^7^. The antigen test’s sensitivity significantly improves when cases with high Ct values (30 - 40) were removed from the analysis ^24, 32^., and this was also proven by our study as shown in table 4.

In order to implement the use of point of care (POC) rapid antigen testing in clinics as well as by the public, we need to improve the efficacy of the test by testing and implementing a scheme that would limit the number of false negative cases, especially in symptomatic patients. We believe that increasing the frequency of the test can improve its diagnostic accuracy. As seen in the sample of 22 patients who had false results by the antigen test in our study, the repeated test showed accurate results in 77% of the repeated test. We have proposed an algorithm that can further improve the diagnostic accuracy of the test in symptomatic patients. The RADT must not be used if more than 7 days have passed since symptoms onset or if the patient has severe symptoms. If the RADT was negative, then the patient must repeat the test again to confirm a negative result by the antigen test. Any positive RADT necessitate further testing by PCR to confirm the infection. This algorithm however should be examined to understand the value of repeating an antigen test in those cases. Moreover, the time frame to repeat the RADT has to be experimented to better understand an appropriate time range to improve the efficacy of the scheme. The definition of mild symptoms must be clearly understood by the public. If a person had severe symptoms or was a high risk individual (close contact), they must perform a RT-PCR as these are higher risk populations and they require a more accurate diagnosis. It is necessary to keep in mind that PCR test is the gold standard test for COVID 19 and that the antigen test can be used in addition to PCR as part of the testing strategies for COVID 19. The use of antigen test can potentially decrease the use of PCR test.

Given the high specificity shown by the RADT, we believe that it can be adequately used in asymptomatic individuals who are not close contacts. The RADT can be used in different settings (gatherings, schools, workplaces) to conduct frequent monitoring of the population and help in identifying cases early to prevent an outbreak. However, its diagnostic accuracy in these settings has to be examined to determine its efficacy.

The study has several strengths. The large sample size and the comparison of nasal swab tested by RADT to Nasopharyngeal samples tested by PCR are the two main strengths in this study. Moreover the use of a single large testing centre allowed standardization and increased quality in sample and data collection. All nasopharyngeal samples were transported and tested in a single lab using the same kits and machines and hence standardizing the results and Ct values. The study has its limitations, the nasal sample was collected using nasopharyngeal swabs. Nasopharyngeal swabs are flexible and smaller and hence are more difficult to collect nasal samples. Therefore, this could have underestimated the results of the study. Moreover, the clinical symptoms of the participants were not collected and there were significant amounts of missing data on time from symptom onset. This had led to a decrease in sample size when testing different models based on restriction of time from symptom onset. This can either under or overestimate the results for the restricted models. Only a small number of cases agreed to have a repeated test after discrepancy in PCR and RADT results. The timing of the repeat test ranged from 24 to 72hrs and wasn’t standardized due to logistical difficulties.

## Conclusion

The sensitivity of rapid antigen test is affected by numerous factors including the viral load, days of symptoms, route of sample collection, and the test mode. The results of the diagnostic assessment of nasal swabs in the RADT used in our study are promising regarding the potential benefit of using them as a screening tool in mildly symptomatic patients. The diagnostic ability was especially high in cases with high viral load. Further investigations ought to be performed to test the algorithms of repeated testing using RADT to further improve its diagnostic ability. More research is required to assess the ability of the RADT to screen large populations with low disease prevalence. It is also important to note that the rapid antigen test is intended to be used alongside RT-PCR and not replace it. RADT can be of benefit in reducing the use of PCR.

## Data Availability

Available upon reasonable request

## DECLARATIONS

### Ethics approval and consent to participate

The study was approved by the National COVID-19 Research and Ethics Committee.

### Consent for publication

All authors gave their consent for publication.

### Availability of data and materials

All the data for this study will be made available upon reasonable request to the corresponding author.

### Author contributions

AA and FM analysed the data and wrote the manuscript. QA, BA and AIA performed data collection. AA and MQ interpreted data and edited the manuscript. MA supervised data collection, data analysis and edited the manuscript. All authors reviewed and approved the final version of the manuscript. Manaf Alqahtani is the guarantor of this work.

